# Effect of Balloon-Blowing on Dyspnea and Oxygenation in Noncritical Adult Covid19 Patients: A Pilot Study

**DOI:** 10.1101/2021.07.27.21260398

**Authors:** Mohammad Bargahi, Soheil Soltani, Nafiseh Rastgoo, Farzane Aryanejad, Roomina Nemati, Mehdi Ghaebi, Arezoo Bajelan, Sohrab Esmaielzade

**Author notes:** This study was registered at the Iranian Registry of Clinical Trial. (IRCT registration number: IRCT20201012049010N1).

## Abstract

Dyspnea and decreased O2 saturation are the most common causes of hospitalization in noncritical covid-19 patients. Breathing exercises and chest physiotherapy are used for managing the patients. These treatments are however not well supported by scientific evidence. In a randomized controlled trial, 80 patients were randomly assigned to planned breathing-exercise (n=40) and control groups (n=40). The participants in the intervention group were instructed to blow into a balloon five times a day while lying down. Other therapies were similar in both groups. The severity of dyspnea at rest/after activity and peripheral oxygen saturation (SpO2) with/without O2 therapy were compared between the two groups on the first, second, and third days. The study findings showed no statistically significant difference in SpO2 with/without O2 therapy on the first, second, and third days between the two groups. Although the severity of dyspnea showed no significant difference between the two groups, the mean score of dyspnea at rest (2.72±2.25 vs. 1.6±1.21, p=0.007) and after activity (4.53±2.04 vs. 3.52±1.66, P=0.017) improved in the intervention group on the third day. Balloon-blowing exercise improves dyspnea in noncritical Covid-19 patients, but it does not significantly improve oxygenation.

## Introduction

Severe Acute Respiratory Syndrome Coronavirus–2 (SARS-CoV-2) has been designated as the cause of a highly infectious pandemic called Coronavirus Disease 2019 (COVID-19), which is spread mainly through respiratory droplets and close contact. ^(1)^ The presentations of COVID-19 are highly variable, with most of the patients (80%) experiencing mild/no symptoms and a minority suffering from pneumonia, severe respiratory failure, acute respiratory distress syndrome (ARDS), or even death (3-5%). ^(2) (3)^

SARS-CoV-2 attacks the respiratory epithelial cells, causing cough, dyspnea, and fever. It induces exudates of serous fluid, fibrin, and hyaline membrane formation within the alveoli. ^(4)^ A chest CT-scan may show varying patterns of lung involvement including bilateral, multilobar ground glass opacification (GGO) with a peripheral/posterior distribution mainly in the lower lobes. ^(5)^ Parenchymal pathological alterations in the form of exudative diffuse alveolar damage are also seen with different degrees of extension. ^(6)^ These pathologies can cause profound hypoxemia with near normal arterial carbon dioxide levels due to ventilation/perfusion (*V/Q*) mismatch and unventilated lung units. The inability to gas exchange often resembles typical ARDS as the disease progresses.^(2)^ Additionally, acute lung injury elicits systemic inflammation and increases pro-inflammatory cytokine expression. ^(2)(7)^

About 20% of COVID-19 patients need hospitalization. One third (32.8%) of the patients experience ARDS during their hospital stay. ^(8)^ Studies have shown that early pulmonary rehabilitation (PR) interventions within two days of admission can reduce mortality in patients with community-acquired pneumonia and interstitial pneumonia. ^(9)^ The primary objectives of PR in this stage are to promote airway clearance and prevent complications of acute illness-related immobilization. Proper incorporation of PR into medical treatment could promote effective expectoration, facilitate mucus clearance, and mobilize secretions to the upper airways thereby improving lung volumes, perfusion, and oxygenation. ^(9) (10) (11)^ In addition, PR may help to prevent or mitigate sequelae related to bed rest, thus improving physical function and outcomes and reducing the length of hospital stay by increasing ventilator free days. ^(9) (4)^

Although there is still no evidence about the efficacy of PR in the specific setting of COVID-19, several established physiotherapy techniques are safely recommended to improve outcomes. ^(9)(10)^ PR has different techniques including breathing exercises (BE) and chest physiotherapy. Balloon-Blowing Exercise (BBE) is a simple, cost-effective, non-personnel dependent BE that can improve lung capacities and respiratory function (12) in people with chronic obstructive pulmonary disease and lower respiratory tract disorders and elderly smokers. ^(13)(14)(15)^

To the best of our knowledge, this is the first trial of the use of a specific BE in covid-19 patients in the acute setting. This study was conducted to evaluate the effects of BBE on dyspnea and oxygenation in covid-19 patients in the acute phase.

## Method

The **Participants** in this study were selected from patients admitted to the non-intensive care unit (ICU) of Booali-Sina Educational and Medical Hospital, Qazvin, Iran, which was designated for covid-19 patients by the relevant governmental committee, from August 15, 2020 to October 31, 2020.

### Inclusion criteria

1) age ≥ 18 year, 2) definitive covid-19 diagnosis (RT-PCR+ / chest CT +), 3) first day of admission in the ward, 4) presence of dyspnea according to the patient’s statement, 5) peripheral oxygen saturation (SpO2) <94%.

### Exclusion criteria

1) pregnancy, 2) history of lung disease (under treatment), kidney disease (under treatment), heart disease, or neurological disease, 3) allergy to latex or balloon material, 4) contraindication to intense aerobic activity by a physician, and 5) need for hospitalization in the ICU/CCU based on the diagnostic and treatment flowchart of the Ministry of Health, 7^th^ edition ^(16)^.

**This study** had a randomized controlled design. The patients who met the inclusion criteria were selected and divided into two groups included intervention and control groups based on random number blocks designed by Excel software. Both groups received the same treatments and oxygen therapy according to the flowchart ^(16)^.

The intervention group was instructed to blow into a balloon at least five sets a day each consisting of five repetitions of BBE in the supine position. They were instructed to inflate the balloon, rest as long as they required, and then blow it again for each set. For BBE, the patients were asked to take a deep breath for 2-5 seconds, hold it in for two seconds, and then inflate the balloon as many times as they could. As a substitute for the balloon when the patient was unable to inflate it, a latex glove size six was used. The balloons and latex gloves were replaced every day. Eleven-inch oval rubber balloons were used.

### Assessment

The patients were assessed on days 1 (before the intervention), 2 and 3. The following data were collected:

1. The participants were evaluated for dyspnea severity using The Modified 0-10 Borg Dyspnea Scale (MBS) every day. All patients received this scale in textual and visual forms. After sitting for five minutes (resting) and walking 50 meters while wearing a finger pulse oximeter (activity), the patients were asked to rate the severity of their dyspnea on a scale of 0 to 10. Santamedical Generation 2 Fingertip Oximeter (Gurin Products) was used.
2. SpO2 was measured during oxygen therapy and after five minutes without oxygen therapy using the above pulse oximeter. In both conditions, the pulse oximeter was worn on the finger and the SpO2 value was recorded after two minutes.

### Study termination

The criteria for study termination were a SpO2 drop of more than 15%, a deterioration of more than 20% in the severity of dyspnea, and need for treatment regimen change according to the medical team.

### Statistical analysis

The collected data were analyzed using the IBM SPSS software version 26 in two unlabeled groups. This pilot study was conducted to estimate the sample size needed for future studies due to lack of data and the innovation of the method used. Unpaired t-test was used to analyze all variables between the two groups. One-way ANOVA test was applied to analyze the variables of each group between various days. Statistical significance was set at ≤ 0.05.

### Ethical considerations

The Ethics and Research Committee of Qazvin University of Medical Sciences approved this study (IR.QUMS.REC.1399.043). Informed written consent was obtained from all participants following a detailed explanation of the examination and study procedures.

## RESULTS

Eighty-six patients were selected of whom three patients refused to participate in the study. Two patients were excluded from the intervention group due to their reluctance to continue the evaluations. One patient was also excluded from the control group after the second day as a result of a change in his treatment regimen. Finally, 80 patients, including 49 men and 31 women, completed the study. **(Fig.1)**

**Fig.1.**
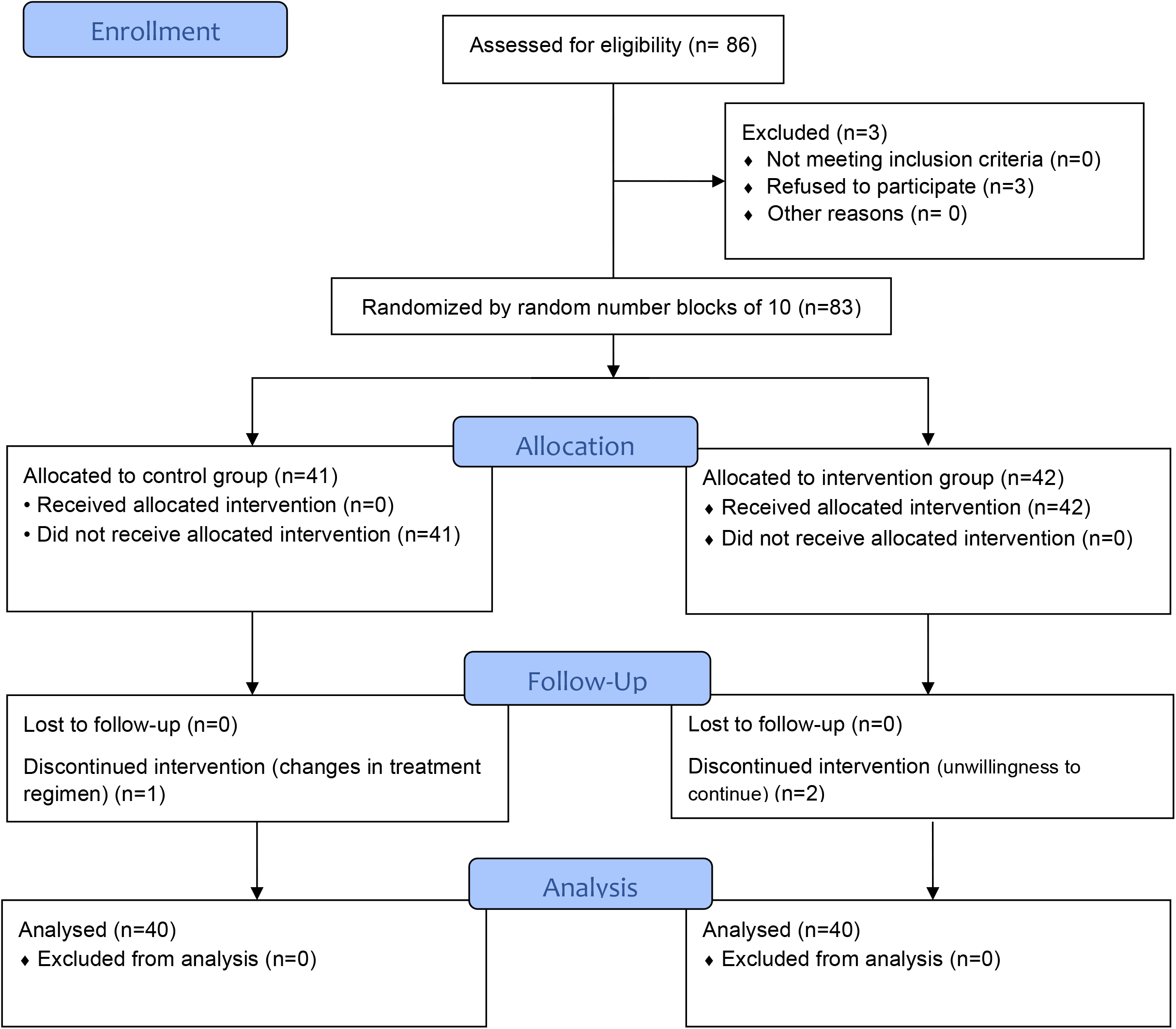
Flow diagram of study ^(20)^

The patients were randomly assigned to control and intervention groups. Both groups received the same drug treatment regimen determined by the relevant medical team. There was no statistically significant difference in gender, age, body mass index (BMI), and extent of pulmonary involvement on CT scan between the groups. **(Table.1)**

**Table1.** Comparison of Demographic characteristics between intervention and control groups

| variable | Control | Intervention | P-value |
| --- | --- | --- | --- |
| Gender, N (%) | M 25(62.6) | M 24 (60) | 0.83 |
|  | F 15(37.5) | F 16 (40) |  |
| Age, yrs (M±SD) | 57.1±18.7 | 58±17.13 | 0.82 |
| BMI, kg/m2 (M±SD) | 26.85±3.22 | 26.81±3.14 | 0.96 |
| **Lung lesions-CT** |  |  |  |
| Multilobar, n (%) | 25(62.5) | 26(65) | 0.82 |
| Unilobar, n (%) | 15(37.5) | 14(35) |  |

M=male, F=female, BMI=body mass index, CT=computed tomography

SpO2 values were compared between the two groups. Twelve participants (15%) showed no increase in the SpO2 value without O2 therapy after three days, of whom four (5%) were in the intervention group and eight (10%) in the control group. However, both groups showed relative improvement on days two and three. Nevertheless, there was no statistically significant difference between the two groups on the first to third day (p>0.05). The SpO2 levels with/without O2 therapy after three days were better in the intervention group compared to the control group (2.8±4.8 vs. 2.8±2.7, and 4.8±4.3 vs. 3.2±2.7), but the differences were not statistically significant (p>0.05). **(Table.2, Fig.2)**

**Table2.**
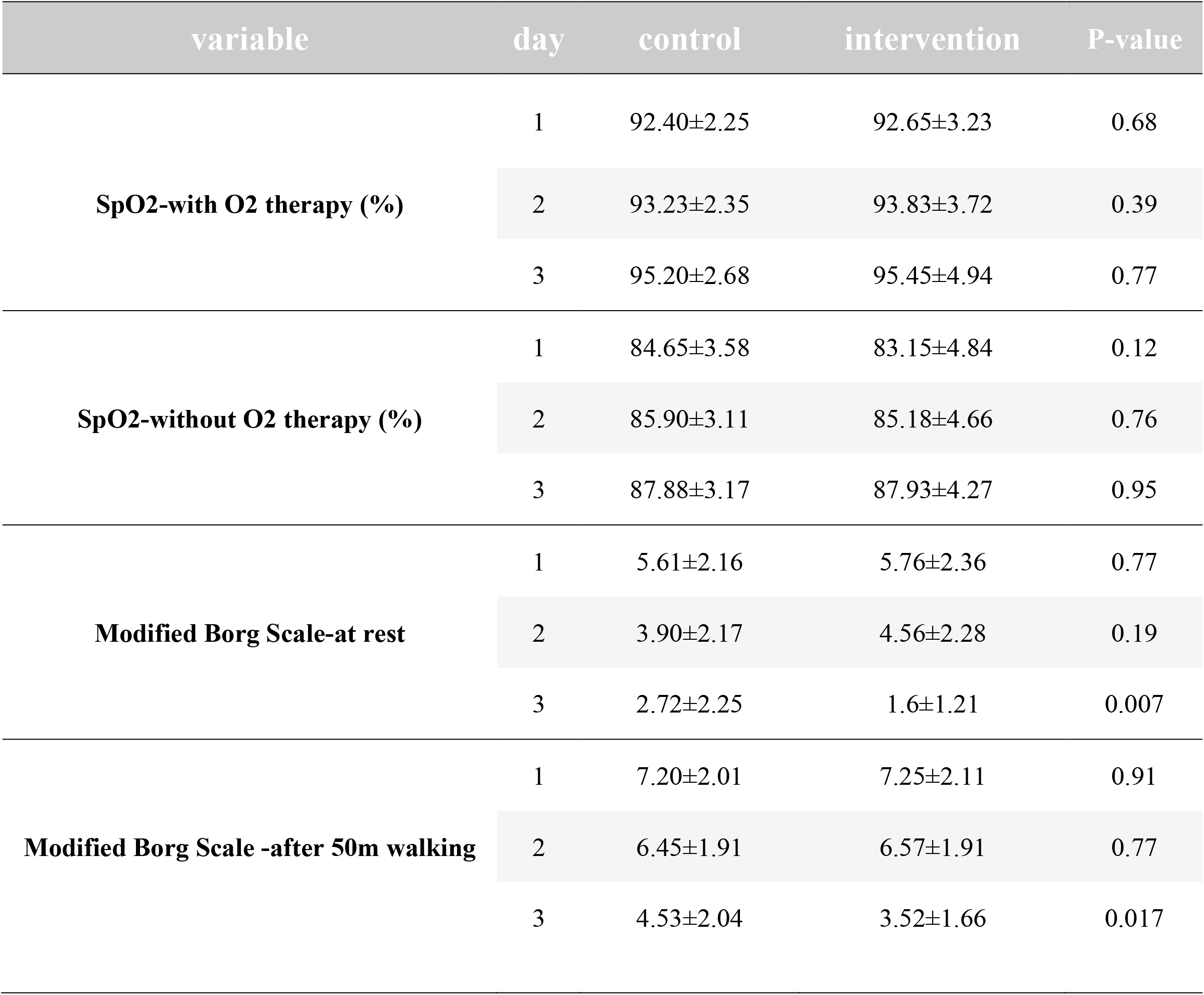
Comparison of Modified 0-10 Borg Dyspnea Scale and peripheral oxygen saturation (SpO_2_) between the two groups (M ± SD)

**Fig.2.**
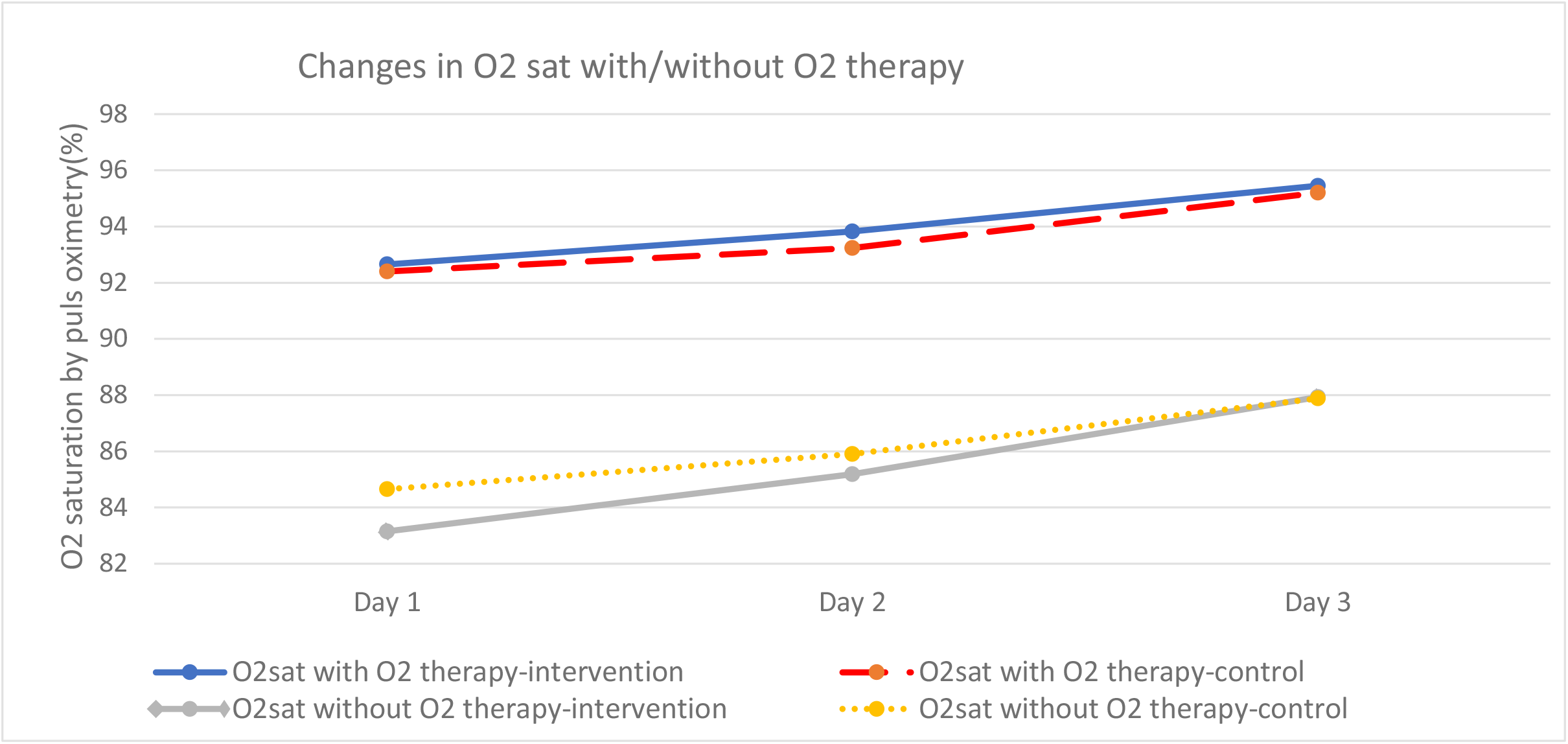
Changes in O2-sat level with and without O2 therapy during the study

There was no significant difference in dyspnea at rest or after activity between the two groups based on MBS on the first day. Breathing improved in both groups on the second and third day; there was no significant difference on the second day (p>0.05) while a significant difference was found between the BBE and the control group (at rest and after activity) on the third day of the trial (p≤0.05). **(Table.2, Fig.3)**

**Fig.3.**
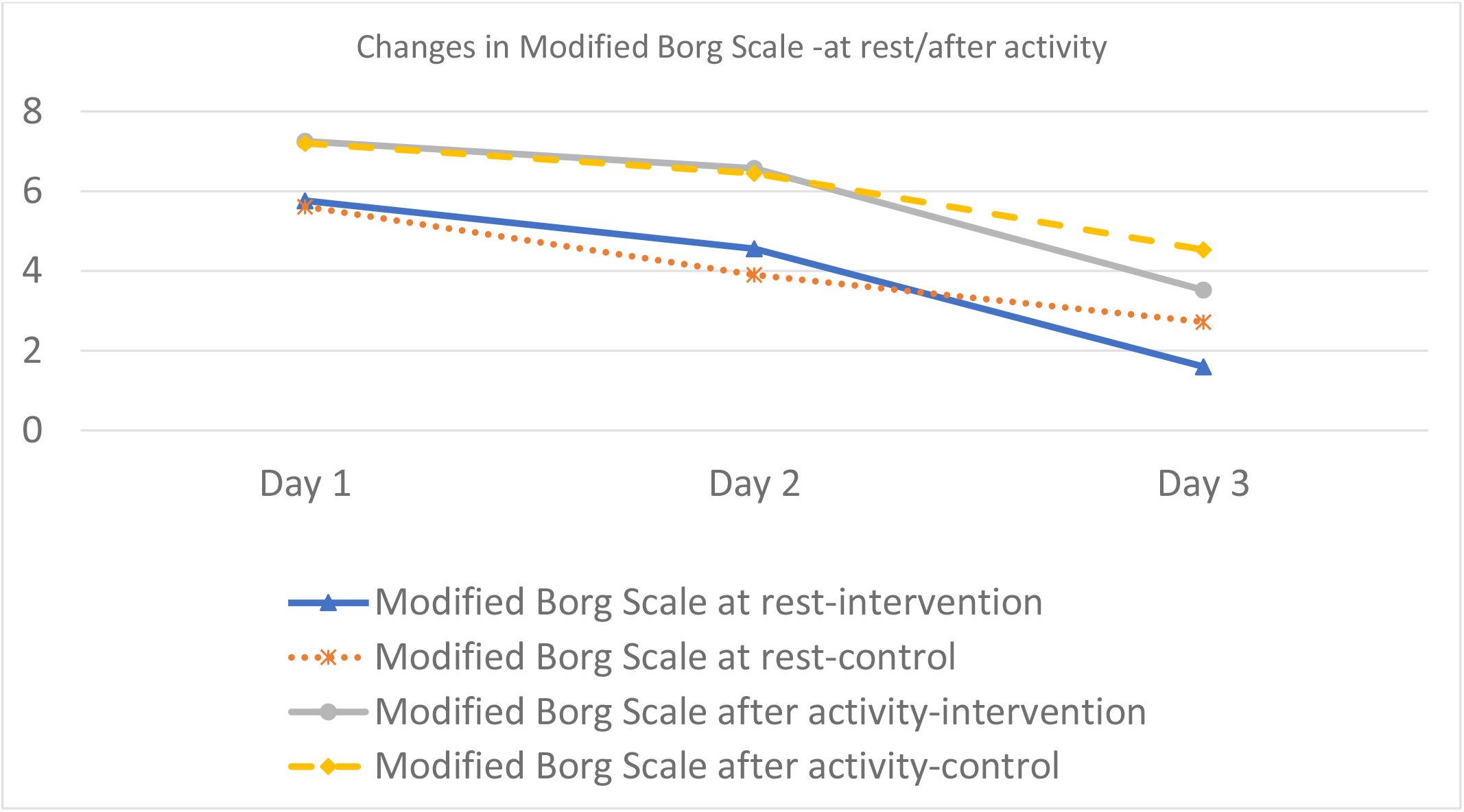
Changes in Modified 0-10 Borg Scale at rest and after activity during the study

Additionally, the participants in case and control groups were divided into four subgroups based on their SpO2 value without O2 therapy on the first day for a more accurate assessment, including SG1 (SpO2<80%), SG2 (80%≤SpO2<85%), SG3 (85%≤ SpO2<90%), and SG4 (SpO2≤ 90%). **(Table.3)** All intervention subgroups experienced an increase in SpO2 levels on the second and third days except for SG4 that had a decrease on the third day compared to the second day. The changes in SpO2 levels in the intervention subgroups SG1 to SG4 were 4.2, 2.3, 2.6 and 1.6 with O2 therapy and 8.8, 4.2, 2.6 and 3 without O2 therapy. **(Fig.4)** After three days of BBE, there was no statistically significant difference in the SpO2 level change with/without O2 therapy between the same subgroups in the control and intervention groups (p>0.05). **(Table.3, Fig.4)**

**Table3.** Comparison of peripheral oxygen saturation (SpO2) between subgroups (M ± SD)

| Subgroups | Number |  | SpO2 with O2-therapy |  |  | SpO2 without O2-therapy |  |  |
| --- | --- | --- | --- | --- | --- | --- | --- | --- |
|  |  |  | pre | post | p-value | pre | post | p-value |
| SG.1 | intervention(n=10) | M=5<br>F=5 | 89.8± 2.74 | 94± 6.31 | 0.89* | 76.7± 2.75 | 85.5± 3.54 | 0.34* |
|  | control(n=3) | M=2<br>F=1 | 88.33± 1.53 | 93± 2 |  | 79± 0.0 | 85± 3.46 |  |
| SG.2 | intervention(n=15) | M=9<br>F=6 | 92.4± 2.99 | 94.73± 5.81 | 0.75* | 82.47±1.3 | 86.67± 4.3 | 0.98* |
|  | control(n=18) | M=9<br>F=9 | 92.7± 1.97 | 94.5 ± 2.71 |  | 82.11± 1.45 | 86.33± 2.91 |  |
| SG.3 | intervention(n=13) | M=6<br>F=7 | 94.77± 2.42 | 97.23± 1.92 | 0.88* | 87.77± 1.67 | 90.38± 3.1 | 0.58* |
|  | control(n=15) | M=7<br>F=8 | 93.53± 1.36 | 96.13± 2.5 |  | 87.4± 1.29 | 89.47± 2.1 |  |
| SG.4 | intervention(n=2) | M=0<br>F=2 | 95± 1.14 | 96.5± 2.12 | 0.86* | 90.5± 0.7 | 93.5± 2.12 | 0.42* |
|  | control(n=4) | M=2<br>F=2 | 94.5± 1.29 | 96.5± 2.4 |  | 90± 0.0 | 91± 2.45 |  |

\*Comparison of O2-sat level changes after three days of intervention with the same subgroup.
M=male, F=female

**Fig.4.**
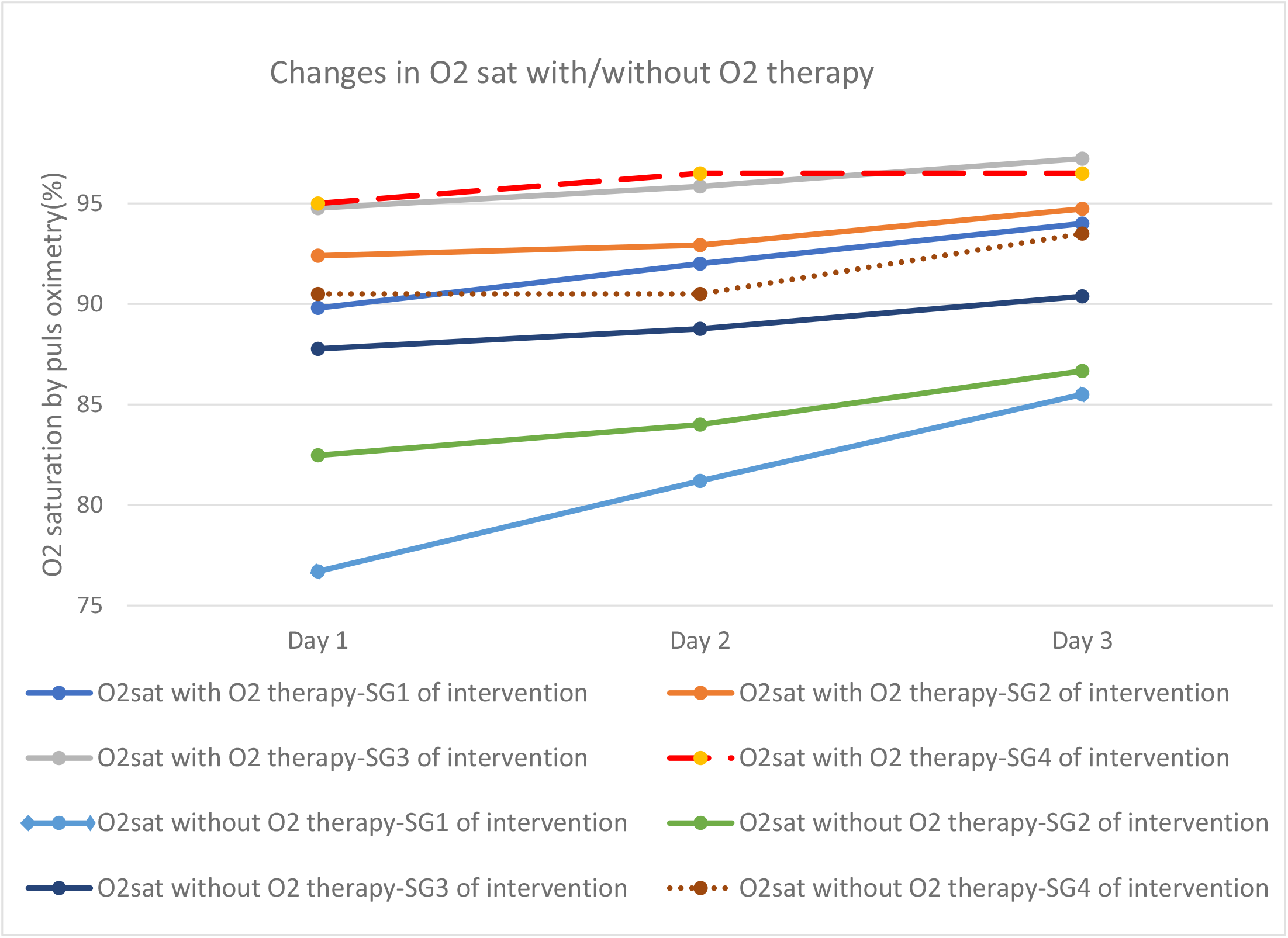
Changes in O2-sat level with and without O2 therapy among intervention subgroups during the study

## Discussion

Dyspnea and decreased SpO2 are the most common causes of hospitalization in covid-19 patients. As mentioned earlier, 20% of covid-19 patients need to be hospitalized due to oxygen-equiring lung infection and one third develop a severe form of the disease. ^(3)(8)^ The most important causes of disease progression may be long-term O2 therapy injuries as well as V/Q mismatch. ^(2)(6)^ Studies have shown that early PR can reduce mortality by clearing the airways as well as improving lung capacity and thus improving gas exchange in patients with interstitial pneumonia. ^(9)(10)(11)^ In a study by Smita Manjusha Das (2018) et al., children aged 3 to 12 years were randomized to receive either balloon or bubble therapy for six days. Among 60 participants, 29 had pneumonia, 17 had bronchitis, and 14 had bronchiolitis. The study reported a significant improvement in SpO2 after regular balloon/bubble inflation. ^(17)^ This finding is in contrast to our study where the BBE group did not show a significant improvement in SpO2 level with/without O2 therapy after three days of BBE compared to the control group. A possible explanation is the difference in BBE days.

In the present study, the severity of dyspnea at rest/after activity reduced significantly after three days of BBE in the intervention group compared to the control group. These findings were consistent with the results of a study conducted by Renuka K (2015) et al. on the respiratory status of patients with lower respiratory tract disorders. In that study, 20 patients received balloon therapy for about 14 consecutive days. The dyspnea scale was measured as a pre/post-test. The study reported a significant reduction in dyspnea after regular balloon inflation. ^(18)^ Combined with our data, it is suggested that the use of BBE is useful in reducing the dyspnea severity in the early days of diagnosis.

There is still controversy about PR strategies in the acute phase of the covid-19. Since BE increases the work of breathing muscles and blood oxygenation alteration leads to a rapid and shallow respiratory pattern, Marta Lazzeri (2020) et al. recommended avoiding such a procedure in the acute setting. ^(19)^ None of our BBE participants met the termination criteria. In the present study, most of the patients in the BBE group reported mild dizziness and difficulty inflating the balloon on the second day. Almost all the patients reported an improvement in their ability to breathe and felt stronger in their chest on the third day. The balloons were not replaced with latex gloves in any of the patients. According to the results, BBE can be done safely in patients with covid-19 in the acute phase. Furthermore, it is highly recommended that patients undergo regular monitoring during BBE.

This study had several limitations. Because of the nature of rehabilitation and assessment, neither the executors nor the participants could be blinded. Therefore, it is not possible to rule out the placebo effect, observer bias or experimenter bias in the current study. In addition, the subjects were limited to patients aged 18 years and above, the duration of the intervention was only three days, and the number of participants was insufficient in each group. Eventually, we did not measure other variables such as the respiratory rate, pulse rate, lung capacities, and anxiety score. Therefore, future studies should address these limitations.

## Conclusions

According to the present study, BBE is a safe medical intervention in noncritical covid-19 patients. It did not significantly improve oxygenation in noncritical covid-19 patients after three days, but it reduced the severity of dyspnea.

## Data Availability

After the preprint is submitted, the collected data will be available by email to the corresponded author.

## Contributors

M. B. conceived, designed, and wrote the manuscript. M. B., N. R., F. A., S. E., and A. B. BBE guidance, data collection, and follow-up of patients. M. B., N. R., and M.G. entered and analyzed data. S. S., M. B, and R. N. edited and reviewed the manuscript. S. S., and M. B. directed and managed the planning and execution of the project. All authors reviewed and approved the final version of the manuscript.

